# The technological innovation and tuberculosis elimination: a Technology Foresight study

**DOI:** 10.1101/2023.04.06.23288235

**Authors:** Roseli Monteiro da Silva, Afranio Kristki, Bernardo Pereira Cabral, Martha Oliveira

## Abstract

In the present study, tuberculosis specialists were surveyed to rate the most effective strategies to eliminate TB as a public health problem by 2050. Then were investigated the most promising emerging technologies for the prevention, diagnosis and treatment of tuberculosis (TB) expected to reach the market by 2035. This Technology Foresight study was specifically carried out by means of a web survey closed questionnaire, which was sent to 29,988 TB specialists worldwide. Of these, 2,657 answers were obtained and analysed. Respondents had demonstrated a high level of academic training (PhD), more than 10 years of professional experience, and a great diversity of both areas of knowledge and geographic reach. In the view of experts, the strategies with the greatest potential impact on epidemic TB were a) shorter time between diagnosis and start of treatment of DS and MDR-TB; b) strengthening tuberculosis control actions in the most vulnerable populations; c) shorter and less expensive regimens for drug resistant MDR/XDR-TB. Regarding the strategies with the highest potential for eliminating TB, our data suggests that the biomedical paradigm is the strongest among the specialists. The most promising technologies expected to reach the market by 2035 selected by the specialists were: (1) new drugs of known chemical classes or new chemical classes; (2) new point-of-care diagnostic tests for DS-TB, drug resistant or multidrug resistant (MDR/XDR)-TB and TB Infection (TBI). We contribute by discussing the most promising technologies and strategies for the elimination of TB in light of social determinants of health models and forecasting studies. We conclude by suggesting that the expected emerging technologies ongoing development will not suffice to end TB by 2050.

## Introduction

Tuberculosis (TB) is considered one of today’s major public health challenges (1). Despite diagnosis and treatment strategies undertaken in the last decades, a slow decline in incidence has been observed globally and is estimated to have decreased 1.7% per year for the period from 2000 to 2019 (2). In light of this statistic, new technologies are indispensable for decreasing TB incidence and eliminating TB as a public health problem (3–4).

According to the Institute for Health Metrics and Evaluation (IHME), the decrease in the incidence of TB worldwide has slowed. Considering the HIV-negative population, the annual rate of reduction of incidence (IRR) decreased from (−1.6%) per year during 1990 to 2005 to (−1.3) during 2006 to 2016 (5,6).

Given this backdrop, the End TB Strategy (8) with the ultimate goal of eliminating TB as a public health problem by 2050 has become increasingly distant. According to Dye et al. (2013), achieving this goal would require, an incidence reduction rate of around 20% per year (7)

To understand the underlying mechanisms by which the disease has remained globally and epidemiologically relevant, tuberculosis (TB) experts have recurrently identified several causal elements such as: social inequality, poverty and consequent social vulnerability (9); the epidemiological transition, the increase of chronic diseases and the subsequent increase of susceptibility to TB (10); the high prevalence of tuberculosis infection (TBI) and increased risk of disease reactivation (11–13); the difficulty of accessing the health system (14) and the poor quality of services (15). Finally, another critical factor that has been cited is that healthcare interventions have been rendered technologically obsolete (16, 5).

Though the literature has cited the need for technological innovations as interventions for TB, there appears to be no consensual agreement regarding specific research, technological, or innovation foci to target. In fact, the literature suggests at least three different lines of thought regarding the relevance and urgency of new technologies or which technologies should be prioritized for the elimination of tuberculosis.

According to the first line of thought, improving access to and quality of care should lie at the forefront, especially in countries with high TB burden. This means that the strategic focus for fighting TB must be on diagnosis and treatment, with emphasis on universal access to the available technological resources (9,14). Such interventions would accelerate the drop of the incidence rate from 7 to 10% per year, which was historically achieved in Europe in the late nineteenth and early twentieth centuries (17). This rationale reinforces the biomedical paradigm for confronting the epidemic, complemented by actions of social protection and governmental commitment. Hence, new technologies are not prioritized in the present and are viewed eventually as part of the future vision of improved care performance (18–20).

According to the second line of thought, there is both a need for new technologies as much as for the improvement of individual care. Unlike the first approach, there is a sense of urgency in regards to technological innovation for the elimination of TB (21, 22, 3, 23, 5, 24). In the report of the Global Strategy for TB Research and Innovation (25), researchers advocate the imperative need for new technologies, which is given equal emphasis as the improvement of current interventions for the elimination of TB.

Finally, experts from a third line of approach claim that elimination can only be achieved with new technologies capable of reducing the prevalence of TBI (screening/screening and prognostic tests, new treatments, therapeutic vaccine) and/or a new vaccine(s) capable of preventing disease, targeted at young adults and adolescents (13, 1, 12, 11, 26, 27). It is worth mentioning that since TB is a disease of respiratory transmission, the scientific community has increasingly come to the understanding that the current control actions are insufficient to interrupt the chain of transmission (4).

Against this backdrop, the present study sought, 1) to identify from the viewpoint of TB specialists, the medical-sanitary strategies with the greatest potential impact on the epidemic, regardless of the current status of technological development, 2) to prospect the most promising technologies to reach the market by 2035 and their potential contribution to the elimination of TB by 2050.

## Method

To answer the research questions, a future study was conducted, by means of the methodological approach known as Technology Foresight (TF). Technology Foresight studies have been carried out since the 1990s in a systematic way in several countries, mainly in Europe and Asia, as support programs for long-term governmental decision making on public policies in science and technology (28).

In the healthcare sector, foresight studies have explored future possibilities for therapies that still pose medical challenges, for instance: (a) the future of cancer care (29); (b) new Treatments for cystic fibrosis (CABRAL et al, 2022); (c) future of genetic therapies for rare genetic diseases (BRAGA et al., 2022) (d) probing expert opinions on the future of kidney replacement therapies (CABRAL et al, 2020).

TF, as explained by Haghdoost et al. “… is a process that takes a systematic long-term look at science, technology, economy, and society in the future (6); it also seeks to identify areas of strategic research and emerging technologies, which seem to result in valuable socioeconomic advantages for the society” 30 (2).

Developed by researchers from the Science Policy Research Unit (SPRU) /Sussex University (31–34), TF is based on the assumptions and concepts of innovation economics, represented by neo-Schumpterian principles (35). Within this framework, innovation is conceived as a phenomenon of intrinsically economic nature and the main driver of economic and social development. Accordingly, Linstone (2011)’s historical analysis identifies three eras of technological development and their socioeconomic impact: (a) industrial, (b) information technology, and the molecular era. For the author, the latter has begun in the mid-1990s, characterized by innovation in biotechnology, material sciences and nanotechnology, presenting the biggest opportunity for fighting TB. (36).

In TF, it is assumed that the future cannot be scientifically demonstrated from certain assumptions. The goal is to address the chances of realization and the options for actionable strategies in the present (37). The exercise of anticipation should be backed by explicit consideration of future possibilities and allow for freedom of thinking. By surpassing routine and preconceived barriers, the approach aims to identify points outside the trend curve - sentinel events – which are indicative of changes or opportunities for innovation.

With the intention to capture the perception/vision of innovation agents and other stakeholders about the technological future, TF uses the flexible combination of several social research tools. In this study, we specifically used a closed questionnaire, applied virtually to researchers around the world, through the platform *Survey Monkey*®.

Invited survey participants and their e-mail addresses were obtained from scientific publications indexed in *Web of Science* (WoS), through the *tesaurus* TS = “tuberculosis” or “tuberculoses” or “Koch disease” or “Koch’s disease” or “*mycobacterium tuberculosis” (via Capes*^1^). A total of 43,087 scientific documents and 29,988 researchers were identified for the selected period of 2015 to 2020. Their email addresses were extracted from the scientific publications using a Python script, developed by the Prospective Studies Group of Fiocruz. Invitations were sent as well as answers were collected between October and December of 2020.

The development of the survey instrument involved an extensive literature review, thematically covering: (1) social determinants and epidemiology of TB (46 documents); (2) expectations about the future of the epidemic (28 documents), (3) emerging technologies in vaccines (31 documents), diagnosis (36 documents), and treatment of TB (22 documents). The review included original scientific articles, review articles, scientific letters and editorials, and technical reports from international organizations specialized in TB. The questionnaire was composed followed questions (1) strategies of greatest potential impact for the elimination of tuberculosis, <u>**regardless of their current technological feasibility**</u>; (2) most promising technological solutions to reach the market by 2035; (3 to 7) specific technologies most promising in vaccines, diagnostic tests and treatment, to reach the market by 2035. For these questions, respondents were asked to assign scores from 1 to 5 for the alternatives presented, being (1) the least promising technology and (5) the most promising technology.

All questions highlighted the 15-year timeframe (from 2020 to 2035) for new technologies to reach the market, with the perspective of eliminating TB as a public health problem by 2050.

### Ethics statement

Confidentiality and privacy were described to all participants prior to enrolment in the study. All data was anonymized, without possibility of participant’s identification. Such security is guaranteed by the software itself. All participants provided consent by the enrolment in the study (See supplementary information – Introduction page). Participation was voluntary and was not stimulated by any gains. Participants were free to give up at any time and an information contact was made available for any questions or issues. The application of the questionnaire did not involve any direct contact between researchers and the participants.

### Respondents’ characteristics

Of the 29,988 invited researchers, 2,711 (9.0%) agreed to participate in the survey and answered the questionnaire. We obtained 2,567 valid answers^2^, of which 69.2% were complete.

Regarding the characteristics of the respondents, the vast majority (95.5%) reported having a post-graduate degree, with 76.4% holding a doctorate. Considering areas of knowledge, 35% of respondents reported having knowledge in basic science areas - microbiology, genetics, immunology; 24% in clinical areas - infectiology, pneumology - and 20% in public health and epidemiology.

Regarding professional activity, respondents were mainly affiliated with teaching and research institutions - only 2% reporting working in industry. About 33% reported working in basic research, 17% in public health, 14% in clinical research, 12% in epidemiology, and 6% in translational research. About 2/3 of the respondents declared professional experience of more than 10 years. Finally, regarding geographical origin, 31% reported being from Asia, 23% from Europe, 16% from South America, 15% from Africa, and 14% from North America.

## Results

Regarding the strategies with the greatest potential impact on the epidemic, the highest proportion of grade 5 were attributed to: (1) reducing the time between diagnosis and initiation of treatment (61%), (2) strengthening control actions in vulnerable populations (59.9%), and (3) reducing the cost and time of MDR-TB treatment (50.1%).

Considering that TB has distinct epidemiological characteristics among geographic regions, the data were also consolidated and analysed according to the origin of the researchers.

Small regional differences in the hierarchy of the most valued alternatives were observed. While researchers from the Americas tended to more value strengthening control actions for more vulnerable populations, researchers from other continents rather prioritized reducing the time between diagnosis and treatment initiation. Researchers from Australasia considered reducing the prevalence of TBI alongside reducing the cost and time to treatment of MDR-TB as factors of the greatest potential impact. For researchers from South America, increasing social protection actions was among the three strategies with the greatest potential impact, differing from the other researchers. Another interesting difference was the higher value placed on shorter treatment regimens for TB-DS for researchers from Australasia and Africa. HIV prevention was also of importance for researchers from Africa (Figure 1).

**Figure 1.**
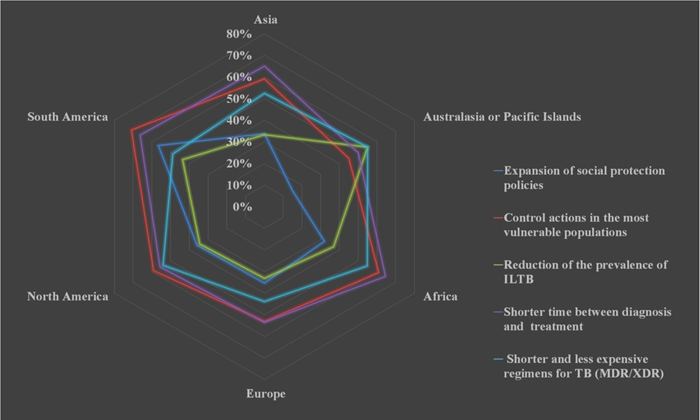
Strategies with the greatest potential impact on tuberculosis, according to the respondent’s origin. Among the most promising technological solutions to reach the market by 2035, new drugs for MDR/XDR treatment (39.6%) and new point-of-care diagnostic tests (39.2%) received the highest ratings. In third place, new drugs or the repositioning of drugs for treating TB-DS (38.3%) were considered most promising. Only 26.3% of respondents considered the possibility of a new vaccine by 2035 as highly promising. Slight variations in the hierarchy of the most promising technological solutions were also observed depending on the origin of the respondent (Figure 2).

**Figure 2.**
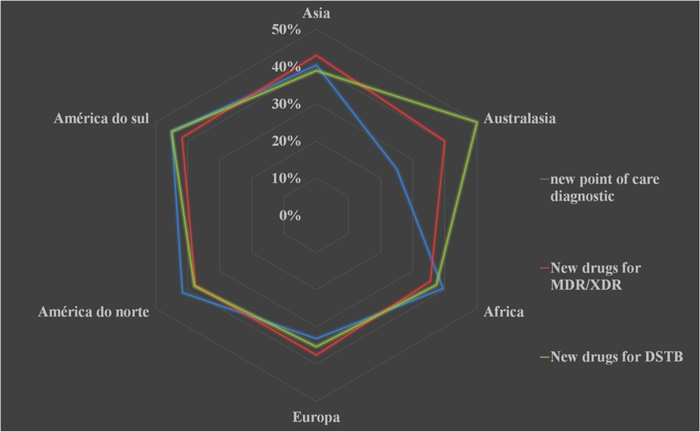
Most promising technological solutions to reach the market by 2035, according to the origin of the respondents. For researchers from Europe and Asia, new drugs for the treatment of MDR/XDR-TB were considered the most promising, while researchers from South America and Australasia considered new drugs for DS-TB as the most promising. Researchers from North America and Africa pointed to new point-of-care diagnostic tests as most likely to reach the market by 2035.

Regarding the specific technological platforms for the development of new TB vaccines, the respondents rated the alternative that assumes an integrated technological platform, capable of combining various techniques and knowledge in vaccinology, as proposed by Zenteno-Cuevas^3^, enabling the complete design of a new vaccine, on new technological bases (26.8%) as most promising (38). This was followed by the adoption of a new route of administration (17.9%), and lastly, a new vaccine using live attenuated organisms inactivated vaccines and recombinant vaccines, including viral vectored (16.7%) (Figure 3).

**Figure 3.**
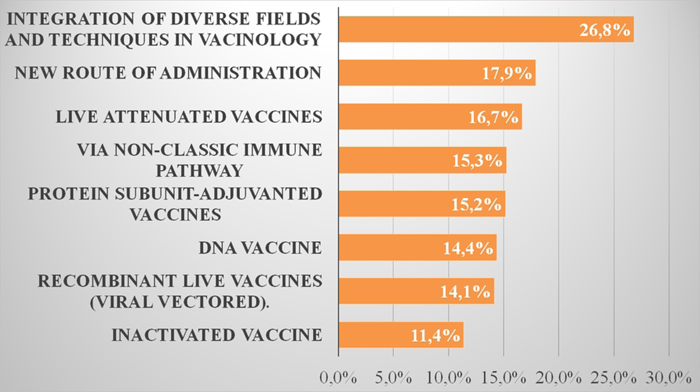
Most promising technological platforms for the development of a tuberculosis vaccine by 2035. Regarding the development of new point-of-care diagnostic tests, the respondents consider the nucleic acid amplification platform (NAAT) by PCR as most promising (38.4%); the new generation of target genomic sequencing (T-NGS) or a whole-genome sequencing (WGS) came in second with 32.9% and in third place the host biomarkers based on “omics technics” (RNA-signature, Micro RNA, metabolomics, proteomics) with 21.1% (Figure 4).

**Figure 4.**
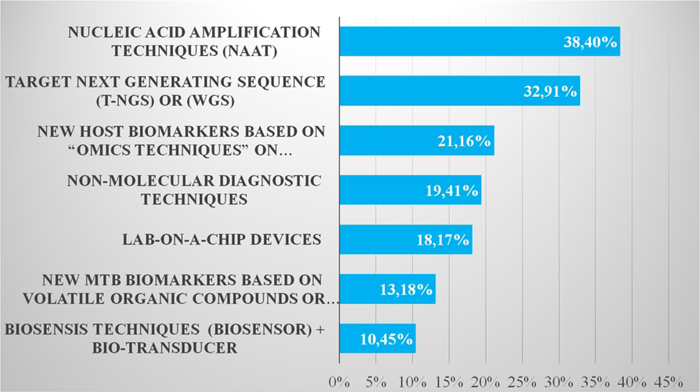
Most promising technological platforms for the development of new point-of-care diagnostic tests for tuberculosis by 2035. Regarding the treatment of all forms of TB, new drugs based on new chemical classes were considered the most promising platform for the treatment of all clinical forms of TB (Table 1). Specifically, for TBI, therapeutic vaccines and immunotherapy with various biological agents (peptides, monoclonal antibodies, interferon). However, for both TB-DS and TB-DR, new drugs from already known chemical classes were considered most promising in second place.

**Table 1.**
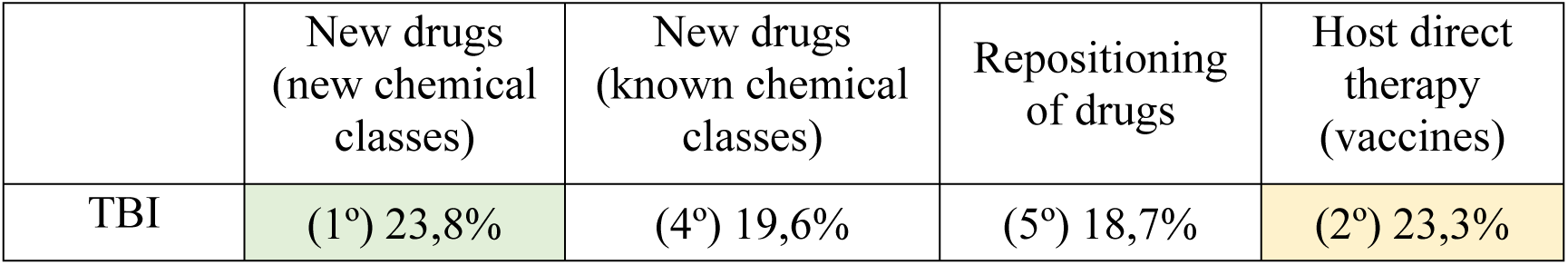
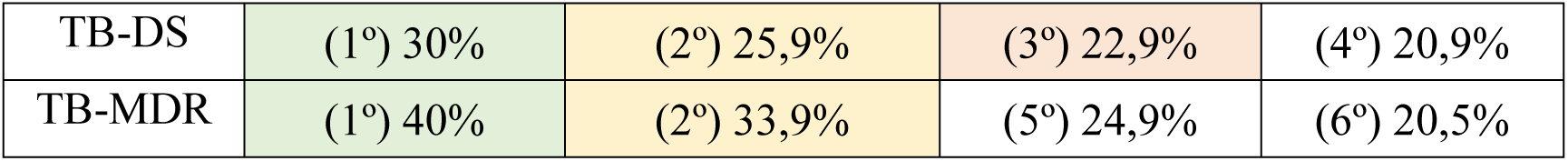
Most promising technology platforms in tuberculosis treatment to reach the market by 2035.

## Discussion

### Strategies with the greatest potential impact for eradicating tuberculosis by 2050

The TB specialists who answered the survey considered strategies oriented towards improving diagnosis and treatment as those with the greatest potential impact on the TB epidemic. Moreover, the intensification of actions targeting the most vulnerable populations was ranked as the second most important factor. It was observed that interventions of a preventive nature - reducing the prevalence of TBI (36.6%), population screening to identify individuals at high risk for the development of TB disease (36.0%) and increasing population immunity (26.8%) - were the least valued strategies.

This response profile reinforce that those researchers are highly driven by the biomedical paradigm. This paradigm is represented by the seminal idea that “TB prevention begins with cure” (STYBLO & BUMGARNER (9). It assumes that the proper diagnosis and treatment of patients (cure), would reduce mortality, prevalence <u>and, on an ongoing basis, incidence</u> (39).

This same paradigm is present in several scientific and technical documents of literature specialized in TB (4). In the guidelines of the End TB Strategy (2015), for example, the first pillar - Patient-Centred: Care and Prevention, specifically expresses more than the importance of care and cure, but additionally emphasizes prevention within appropriate diagnosis and treatment of cases (WHO, 2015). In the same sense, Lönnroth et al. (2009) reinstate that the main challenge of TB control is to ensure globally adequate access to diagnosis and treatment (9). They point out that options to include preventive efforts have not yet been fully considered.

Thus, the priorities selected by the specialists appear to lie within the biomedical paradigm, in line with KUHN’s (1998) premise:

> “The commitments that govern normal science specify not only what sorts of entities the universe does contain, but also, by implication, those that it does not.” <u>40</u> (26)

It is worth emphasizing that the biomedical paradigm does not deny the existence of social factors that interfere with contracting TB. Only, such conditions are treated as externalities and not as part of a complex process of social production. Therefore, it is not paradoxical that the strengthening of control actions aimed at the most socially vulnerable population groups has been listed among the most prioritized actions. Notably, no specific epidemiological studies were found on the potential effects of this strategy, nor on new drugs for the treatment of MDR-TB for the elimination of TB.

Moreover, prospective studies based on Technology Forecast (31), have indicated opposite expectations regarding specialists’ responses about the potential impact of new strategies and technologies for TB elimination.

Dye et al (2013) estimate that reducing the time between diagnosis and the beginning of treatment would have an impact on the order of 28% in reducing the incidence of TB over a 35-year time span (3). Regarding the high prevalence of TBI, the authors point out that even if TB transmission were totally interrupted in 2015, the existing pool of infected persons would still generate more than 110 cases per million population in 2050 (hypothetical case of 1100 new cases per million/year). On the other hand, if only 8% of individuals infected with M. tuberculosis were permanently protected from the disease each year, the incidence could drop to 90 new cases per million. If the intervention reached 14% of those infected, the incidence could drop to 20 cases per million by 2050.

According to Abu-Raddada et al (2009), reducing the treatment time for TB-DS, would also have a limited effect on incidence, with an estimated reduction of 8.3%; 19.3%; 22.9% with shortening of the therapeutic regimen to 4 months, 2 months, and 10 days, respectively (41).. On the other hand, the authors estimate that neonatal vaccination, considering the portfolio of vaccines listed in the study, could decrease the incidence of tuberculosis by 39% to 52% by 2050. They point out that a vaccine targeting adolescents and young adults would have a greater and faster epidemiological impact than vaccination of neonates, an estimated 80% reduction in incidence by 2050 (41).

On the other hand, based on the model of social determinants of health (SDH), derived from the Public Health paradigm, it is possible to unveil an explanatory network, organized in layers of intertwined causal processes, involving from macro elements of the social production of the disease to its “natural” history, at the level of each individual.

It is noteworthy that TB has benefited nowadays from the negative effects on health, stemming from the urbanization process (agglomerations) that favor transmission and from the modern lifestyle (increase in chronic non-communicable diseases, including mental illness and drug addiction) that increase the risk of contracting TB (42).

The slow reduction in the incidence and increase in the proportion of MDR-TB has demonstrated that current control actions are progressively losing effectiveness and that there is a great need for new interventions primarily oriented towards prevention of illness, early detection, and reduction of the pool of infected people.

Schrager et al. (2020) reaffirm that only a new vaccine with the ability to prevent tuberculosis, targeted at adolescents and young adults, could effectively contribute to reducing incidence. If introduced by 2025, such a new vaccine could result in a drop in incidence of nearly 17% per year, effectively paving the way to eradication (43). In addition, for several authors, immunization would be one of the most effective strategies to address bacterial resistance, since the genetic sites of immunity are distinct from those that generate drug resistance (38).

In a prospective epidemiological study on the potential impact of a new vaccine, Harris et al. (2020), estimated that the expected reduction of incidence (IRR) in this period would be 51%, 52%, and 54% respectively for China, South Africa, and India, using 10 years of massive vaccination campaign, directed to adolescent population and an efficacy of 70% in preventing the disease as parameters (26). According to the authors, preventive action targeted only at the infected population would have a greater and faster impact, considering the goal of eradicating the disease by 2050.

On the other hand, in closeness with the public health paradigm, researchers such as Zenteno-Cuevas (2017), Schrager (2018,2020), Keshavjee (2019), Ole F Olesen (2019), Roland Diel (2013), Philipe Glaziou (2013), Holmes (2017); Barry Bloom and Ranif Atun (2013), Helen McShane (2011), advocate the need for new strategies, in particular aimed at reducing the huge contingent of infected (TBI) and preventing the active disease in the general population. However, even today a minority of experts support this view.

A second line of explanation for these results may be related to the tendency observed in TF exercises, that respondents should be captured by current problems, rather than theoretical reflection on future possibilities. The empirical, tangible reality sometimes imposes itself over an abstract, future-oriented logic. Thus, the focus on interventions with long-term epidemiological impact are displaced by the need for short-term care solutions.

Bonaccorsi et al, from the cognitive psychology and decision theory paradigm, poses the possibility of the occurrence of cognitive bias in technology foresight exercises conducted with experts only (44). According to this paradigm, competence to make judgments and decision making processes depend on memory capacity and the nature of the information being processed.

> “People do not perform exhaustive search and analysis of alternatives, but rather use well-functioning (but not optimal), rules of thumb (*heuristics*), and are subject to systematic distortions (*biases*) in their judgments (particularly, probability judgments) and decisions. (44) (2)

The participation of several stakeholders in foresight exercises is a seminal recommendation as a strategy to prevent biases. For technical-operational reasons, in this study, the exercise was conducted only with TB experts.

Aiming to mitigate the occurrence of this type of bias, Bolger and Wright (2017) suggest the application of multiple techniques - a process called expert knowledge elicitation (EKE) that makes it possible to confront, at different analytical stages, information from several sources (45). In this research, forecast studies and literature review data were used as analytical counterpoints to the research findings.

Such limitations, however, do not invalidate the method, nor its results. It only indicates that the future may result from this dominant thinking, in this case, linked to a biomedical view of the problem.

It is important to stress that the specialists’ choices may be able to solve important problems in clinical practice, which, on its own, is remarkable. Nevertheless, bearing in mind the expected reduction in the incidence of TB, we believe that such priorities will have little impact on the elimination of TB.

### Most promising technological solutions to reach the market by 2035

Most respondents reported that point-of-care diagnostic tests and new drugs for MDR-TB would be the most promising technologies to reach the market by 2035. Given the very close scores (39.6% and 39.2%), the results suggested that the experts’ highest innovation expectations were also focused on diagnosis and treatment. New drugs for treating TB-DS was the third technological solution with the greatest potential to enter the market.

There was also a rather skeptical attitude toward the emergence of new TB vaccines and prognostic tests for individuals with TBI. Indeed, as discussed in a later section on new diagnostic technologies, significant technological advances in the last 10 years have made the field quite dynamic (42). NAAT Technology-based diagnostics have incorporated incremental innovations that bring new products closer to the ideals of point-of-care diagnostics and operational simplification (46). A technological path that adds to PCR-based nucleic acid amplification other technologies such as microfluidics, biosensors, and nanotechnology is already feasible.

This phenomenon has not been observed in the vaccine area, except for the use of technological platforms focused on the production of protein subunits, the most used in the development of several vaccine candidates. Other technological platforms such as recombinant or inactivated vaccines have also not been able to open a path of innovations. In the drug area, the most recent technological development also shows promising results with new molecules from new chemical classes. At least 18 new drugs from this category are in phase III of clinical trials (47).

This evidence, in the light of the postulates of the Economics of Innovation, suggests that the resources, the interests of public and private innovation agents, and the processes of interaction and learning have proven to be more effective in the diagnostic area. It is worth remembering, in the neo-Schumpeterian line of thought, that technological innovation is:

1. a continuous and cumulative process, involving not only incremental and radical innovation, but also its diffusion, appropriation, and use (48);
2. a process involving interactive learning and accumulation of knowledge, which takes place in connection with ongoing activities in production and sales (48);
3. institutions, in different innovation systems, affecting the nature of these interactions between people and customs, which, in part, may explain the accumulation or non-accumulation of learning processes (49)
4. intensive learning processes can be converted into technological innovation avenues or technological pathways (49, 50)

Thus, as an economic phenomenon, it is likely that technological innovation in diagnosis has been recognized by innovation agents in industry, governments, research organizations, and the financial system as more timely and promising -all this with the proper scientific backing of the biomedical paradigm.

It should be noted that technology is itself a body of knowledge about certain classes of events and activities, and not a mere application of knowledge brought from another field. The normal situation in the past, and to a considerable degree also in the present, has been that technological knowledge precedes scientific knowledge (e.g. BCG vaccine) (51).

### Most promising vaccine technologies (to 2035)

The technological platform most valued by respondents for developing a new tuberculosis vaccine corresponds to the approach proposed by Zenteno-Cuevas (26.8%), defined as follows:

> This new concept of vaccinology encompasses the utilization and integration of diverse fields and techniques of study, including reverse vaccinology, immunogenomics, immunogenetics, systems biology, immune profiling, proteomics, whole genome sequencing, transcriptomics, host–pathogen interaction, bioinformatics, and computational modelling. 38 (5)

However, the scattering of the values assigned to the listed alternatives was observed. This, even considering the integrated technological platform proposed by Cuevas obtained 8 percentage points above the second option (new routes of administration - 17.9%) and double the score received by the inactivated vaccine technology (11%), considered the least promising of the technologies.

In the Pipeline Report Tuberculosis Vaccines 2021, there are 15 vaccine candidates listed, which are in clinical trials, distributed from the technological point of view in: a) inactivated/fragments (3); b) attenuated MTB (1); c) protein subunits (5); d) viral vector (3), e) BCG (2) (52). No candidates in this portfolio have been developed based on an integrated or mRNA/DNA platform (52).

Mike Frick, in an introductory text to this report, offer an interesting reflection on the dynamics of technological development in the area of tuberculosis vaccines, compared to the results achieved against COVID19.

While the technological development time was measured in months to obtain 20 vaccine candidates during the COVID19 pandemic, the technological development time used for TB – a disease that kills more than 1 million people per year – is measured in centuries.

> Nowhere to be found is the tool that would reduce TB incidence in time to realize the World Health Organization’s vision of eliminating TB by 2030: a new vaccine (or multiple vaccines) that protects adolescents and adults against TB. The confluence of the BCG centenary and the advent of COVID-19 vaccines begs the question: why are there no new vaccines against TB? 52 (1).

The author points out the large volume of resources applied (about 11 times what was applied in TB in the last decade) and the regulatory changes, which enabled the parallel achievement of some stages of technological development as main factors related to the success of this technological innovation process.

In addition, Frick highlights the key role of governments and funders in the proper direction of resources in the promotion of this process. This explanation brings us back to the innovation ecosystem approach, where innovation is the result of the push of demands arising from public health policies into the market. According to LI, GARNSEY (2014) “This requires new sources of funding and new relationships between public sector organizations, philanthropic trusts, medical foundations and entrepreneurial innovators” 53 (1). In the history of technological development of TB vaccines, we highlight some lessons learned:

- Immunity produced by the classical T-lymphocyte induction pathway is not sufficient to produce protection against TB. Vaccines with a single dominant antigen, as a booster to BCG, do not seem to be as effective as expected. This has somewhat called into question technologies based on protein subunits and viral vector (Bloom and Lambert, 2016).
- Bacillus’ containment is a more tangible goal from a technological perspective than preventing primary infection or sterilizing the granuloma (54).
- Adolescents and young adults should be the preferred population group for a new vaccine, considering the goal of TB elimination (26).
- Several vaccine schemes should be considered in the process of developing a new vaccine (55).
- New vaccines are probably the best strategy for tackling MDR-TB (27).

The M72/AS01E (GSK) vaccine, protein subunit-based (containing Mtb32a-Mtb39a dormancy phase antigens), is the closest candidate to reach the market by 2035 (27). The efficacy observed in phase II studies is 54%. This may mean that we are still far away from a new widely effective TB vaccine, in accord to expectation of the respondents.

### Diagnosis and prognosis of tuberculosis

The technologies that had the highest scores from experts for point-of-care TB diagnosis were those based on nucleic acid amplification (38.4%) by polymerase chain reaction (PCR) and new generation of gene sequencing (WGS or T-NGS) (32.9%).

Two issues should be considered when analyzing these results. First, incremental innovations in NAAT technology have been running for more than 20 years, with a number of new products becoming available on the market, including LoC devices that do not require a laboratorial environment or even electrical power (56).

In fact, NAAT is one of the most promising technologies to become an effective point-of-care diagnostic tool, progressively more accessible, simultaneously meeting the demand for diagnosing tuberculosis and mapping the profile of bacterial resistance.

At present, several products are available on the market and arebeing recommended by the WHO: Gene Xpert ULTRA, Truenat MTB plus, Easy NAAT MTBC, FluoroType® MTB (RT-PCR), BD Max MDR-TB (57). The VereMTB Test, in Lab-on-a-chip device by Veredus (Singapore), is under validation within research laboratories (42). The rise in market competition for the new tests should contribute to cost reduction and further diffusion of the technology to poorer countries.

The new generation of gene sequencing (WGS) used to study DNA from isolated *M. tuberculosis* although highly valued by respondents, applies more to Mtb genomic surveillance than to clinical practice. On the other hand, the use of T-NGS in clinical samples has an advantage because other than the potential of miniaturization and low operational cost, with the use of metal-oxide semiconductors and PCR reagents in very few quantities, the use of T-NGS can allow for TB-MDR/XDR diagnosis in 2 to 3 weeks (58).

However, none of these technologies are applicable to the diagnosis of TBI, remaining a serious obstacle to clinical practice and TB epidemiological control measures because they do not extend the diagnostic capacity in people with immunodeficiency, chronic diseases and in children by using sputum as a biological sample (46). Prior TBI testing is recommended for HIV-uninfected contacts over five years of age, as TPT is more likely to benefit those with positive tuberculin skin tests (TST). TST with PPD Rt-23 has low specificity in BCG-vaccinated populations and chain distribution limitations. Interferon-gamma release assays (IGRA) are acceptable alternatives, but the costs and the need for laboratory infrastructure are a matter of concern. New TSTs, using recombinant ESAT6-CFP10 antigen proteins, which include C-Tb (AJVaccines, Denmark, to be transferred to Indian Serum Institute), Diaskin Test® (Diaskin, Generium, Russian Federation), EC-test (Anhui Zhifei Longcom, China) have been developed to increase specificity in laboratory-free settings and recently recommended by WHO (59).

Host molecular biomarkers appear as the third most promising technologies (21.1%). The recent discovery of the ability of these markers to discriminate individuals with active TB from healthy individuals and with TBI (60–62), probably opens new routes of technological development (63).

MacGregor-Fairlie et al. (2020) note that TB diagnosis has been through several transformations, including the use of part of the bacillus (rather than the whole bacillus) in direct diagnosis and, most importantly, the introduction of indirect methods related to the host immune response profile (46).

Nevertheless, these technologies were considered less promising by the respondents. Point-of-care testing has the possibility of radically changing the method of laboratory production, specifically through means of decentralized approach and without a large technical equipment. The central laboratory may be replaced by a chip in the future, distributed throughout the primary health care network.

Two explanatory hypotheses can be raised in relation to these results. First, the medical tradition in the area of TB reinforces the need to identify the bacillus in order to make a diagnosis. Either due to the complexity of the disease or the need for differential diagnosis, it is plausible to assume a bias in this perception. A distorted perception that distinguishes tuberculosis from other infectious diseases with regard to technical requirements to the diagnostic and treatment processes.

The second aspect refers to the relevance given more to the diagnosis of clinically active disease than to the detection of TBI or increased risk of disease (prognosis). The biomedical paradigm prevails in the experts’ expectations.

The use of host biomarkers for the diagnosis of TBI and risk of illness (TBI/TB) could contribute to the control of the epidemic insofar as they make it possible to direct preventive treatment or the use of a therapeutic vaccine. This aspect is highlighted in the WHO report on research and technological development (2020), reiterating the need for increasing investments in this sector (WHO, 2020).

It is crucial to note that such strategic directing would face significant trade barriers due to competition and eventual replacement of NAAT-based technology.

### Treatment of Tuberculosis

Most respondents considered new drugs from new chemical classes as most promising for TB treatment, regardless of the clinical form (TBI or active TB) or the drug resistance status (DS or DR/MDR). In the case of TBI, however, host direct therapy, by means of vaccines, was considered the second most promising option and with very close value to new drugs in new chemical bases.

For TB-DS, drug repositioning still appears as the third most promising technological option. For MDR-TB, new drugs in new chemical classes was the proportionally most valued alternative, with 40% of the responses scoring (5).

Currently there are 30 drugs in various stages of preclinical and clinical development (47). Of this list, twelve are new drugs from known chemical classes, with or without a lead drug. Eighteen constitute new chemical structures with new mechanisms of action. Of the drugs in phase 2-3 of technology development, seven are new drugs from new chemical classes and three from known chemical classes, which demonstrates the strength of the technology development arena^4^.

These data support the experts’ choice of drugs from new chemical classes as the most promising solution for the treatment of TB. For TBI, the immunotherapy strategy through vaccines still constitutes a technological challenge.

## Conclusion

The biomedical paradigm constitutes the main anchor that sustains current strategies of intervention in the TB epidemic. This thinking has been reflected in the prioritization of new strategies, which still prioritize a future focused on diagnosis and treatment as having the greatest potential impact on the control and eventual elimination of TB.

Strategies and technologies aimed at preventing TB illness (preventive vaccines) and eliminating the TB contingent (diagnostic and prognostic tests, preventive treatment and/or immunotherapy) were the least valued by respondents, reiterating the dominance of biomedical thinking among experts.

The technological solutions considered most promising also focus on TB diagnosis and treatment - new diagnostic tests and new drugs for MDR TB. Given the likely materialization of this forward-looking vision, it is possible that new technologies will have little or no effect on the elimination of TB as a public health problem by 2050.

Technological developments in new vaccine development show a reduction in the use of the recombinant BCG-based platform and an increase in those vaccines based on protein subunits. The most promising technological platform, however, in the respondents’ view would be based on a new paradigm, which comprises the integrated application of various areas of knowledge and techniques in vaccinology. Nevertheless, no vaccine candidates from the Pipeline Report 2021 for clinical trials, have been developed based on this paradigm (52).

The most promising technologies in TB diagnostics do not meet the demands for differential diagnosis (TBI and TB), nor do they have prognostic value on the risk of progression from TBI to clinically active disease (TB), or markers for treatment monitoring. Such elements are considered, from a public health point of view, essential for the elimination of the disease. Once again, the issue of governmental involvement pulling into the market (demand pull) innovations that meet public health demands seems to be fundamental for the development of new products of interest to public policies.

Technological trends recorded in the literature point to host biomarkers as one of the most suitable solutions for point-of-care diagnosis of TB and TBI. However, the economic dynamics for developing this technology do not seem to benefit biomarker area. It is also worth noting that although some of these biomarkers have been available since 2018, there has not been a WHO endorsement recommending this technology for new product development.

In summary the most promising specific technologies in the area of TB vaccines, diagnostic tests and treatment were respectively:

1. an integrated technology platform enabling the application of the various fields of knowledge and techniques in vaccinology, including reverse vaccinology, immunogenomics, immunogenetics, biological systems, immune profiling, proteomics, whole genome sequencing, transcriptomics, bacillus-host interaction, bioinformatics, computational modelling.
2. new diagnostic tests based on nucleic acid amplification technology (NAAT) and next generation gene sequencing (NGS).
3. new drugs from new chemical classes for all forms of TB. For ILTB immunotherapy using vaccines was considered the second most promising technology.

## Recommendations

In light of the study results, we recommend a strategic redirection of tuberculosis R&D efforts toward technologies for which the greatest potential impact on the incidence of tuberculosis is expected, with a view to its elimination as a public health problem by 2050.

To this end, firstly it is deemed necessary to strengthen and disseminate a comprehensive vision of the tuberculosis epidemic, so that the fight against the disease is adequately oriented to its network of causalities, in a contextualized manner.

Such an approach should also enable a greater consensus in the scientific community regarding the role of technological innovation, and priorities in DT and Innovation, as an indispensable element of the effort to eliminate the disease.

From a pragmatic point of view, the results from this study suggest that the resources and efforts of technological development and innovation should be directed to the generation of:

- new diagnostic and prognostic tests that enable the screening of the population with ILTB and identification of those at high risk of developing the disease. From the technological standpoint, this means investing in omics technology platforms aiming at the creation of host biomarkers capable of differentiating the clinical forms of the disease;
- new point-of-care diagnostic tests for tuberculosis also based on omics technology, preferably replacing sputum as the biological sample.
- new preventive vaccines directed especially to the adolescent and young population, infected or not by M. tuberculosis.

Such an approach implies directing resources especially to clinical and translational studies, with emphasis on

- multicentre phase (I, II, and II) clinical trials from the current portfolio of TB vaccine candidates. Although many vaccines are ready for human clinical trial, this stage has not yet been initiated.
- clinical studies on the effect of therapeutic vaccines aimed at increasing efficacy, reducing the time of treatment of MDR-TB or decreasing the number of drugs applied;
- technological development and clinical studies in host molecular biomarkers, with the perspective of building point-of-care devices capable of recognizing the clinical forms of tuberculosis, bacterial resistance, and also providing prognostic vision.

Through these suggested processes, the role of governments in the appropriate allocation of resources for research and innovation becomes more critical.

## Data Availability

the survey database was made available in its entirety through the file named Survey_results file extracted directly from the Survey monkey tool. Data will be available upon request.

## Footnotes

1 CAPES – Coordination for the Improvement of Higher Education Personnel, Ministry of Education of Brazil.

2 Respostas dos especialistas que manifestaram ter conhecimento sobre o objeto da pesquisa

3 This new concept of vaccinology encompasses the utilization and integration of diverse fields and techniques of study, including reverse vaccinology, immunogenomics, immunogenetics, systems biology, immune profiling, proteomics, whole genome sequencing, transcriptomics, host–pathogen interaction, bioinformatics, and computational modeling (p.5).

4 https://www.newtbdrugs.org/pipeline/clinical?field_advancing_value=All, Access on: 08/07/2022

## Notes

### Competing Interest Statement

The authors have declared no competing interest.

### Funding Statement

The research was conducted autonomously, without specific funding. It is the result of research from the PhD Program at the Federal University of Rio de Janeiro Brazil

### Author Declarations

All participants were duly informed about the research and agreed to participate in the research, being guaranteed the confidentiality of the answers and the non-identification of any participant. This was done by means of a letter via e mail and the act of agreement duly registered.

## References

1. Bloom B, Atun R. Back to the future: Rethinking global control of tuberculosis. Sci Transl Med 2016; 8(329):329ps7.

2. WHO. Global tuberculosis report 2020; 2020.

3. Holmes KK, Bertozzi S, Bloom B, Prabhat. J., editors. Disease Control Priorities: Major Infectious Diseases [Capítulo 11 Tuberculosis]. (vol. 6).

4. Pai M, Furin J. Tuberculosis innovations mean little if they cannot save lives. eLife 2017:1–9.

5. GBD Tuberculosis Collaborators. The global burden of tuberculosis: results from the Global Burden of Disease Study 2015. Lancet Infect Dis 2018; 18(3):261–84.

6. GBD Tuberculosis Collaborators. Global, regional, and national burden of tuberculosis, 1990-2016: results from the Global Burden of Diseases, Injuries, and Risk Factors 2016 Study. Lancet Infect Dis 2018; 18(12):1329–49.

7. Dye C, Glaziou P, Floyd K, Raviglione M. Prospects for tuberculosis elimination. Annu Rev Public Health 2013; 34:271–86.

8. WHO. End TB Strategy: Global strategy and targets for tuberculosis prevention, care and control after 2015; 2015.

9. Lonnroth K, Jaramillo E, Williams BG, Dye C, Raviglione M. Drivers of tuberculosis epidemics: The role of risk factors and social determinants. Social Science & Medicine 2009; 2009 (68):2240–6.

10. Divangahi M, editor. The New Paradigm of Immunity to Tuberculosis. New York, NY: Springer New York; 2013. (vol 783).

11. Keshavjee S, Amanullah F, Cattamanchi A, Chaisson R, Dobos KM, Fox GJ et al. Moving toward Tuberculosis Elimination. Critical Issues for Research in Diagnostics and Therapeutics for Tuberculosis Infection. Am J Respir Crit Care Med 2019; 199(5):564–71.

12. Esmail H, Barry CE, Young DB, Wilkinson RJ. The ongoing challenge of latent tuberculosis. Philos Trans R Soc Lond B Biol Sci 2014; 369(1645):20130437.

13. Diel R, Loddenkemper R, Zellweger J-P, Sotgiu G, D’Ambrosio L, Centis R et al. Old ideas to innovate tuberculosis control: preventive treatment to achieve elimination. Eur. Respir J 2013; 42(3):785–801.

14. Burki T. Tuberculosis post-2015: looking to the future with optimism. Lancet Infect Dis 2015; 15(1):21–2.

15. Fullman N, Yearwood J, Solomon M Abay, Cristiana Abbafati, Foad Abd-Allah, Jemal Abdela et al. Measuring performance on the Healthcare Access and Quality Index for 195 countries and territories and selected subnational locations: a systematic analysis from the Global Burden of Disease Study 2016. Lancet 2018; 391(10136):2236–71.

16. Pai M, Memish ZA. New tuberculosis tools are here: can we deliver them for maximal impact? J Epidemiol Glob Health 2013; 3(1):1–2.

17. Lönnroth K, Castro KG, Chakaya JM, Chauhan LS, Floyd K, Glaziou P, et al. Tuberculosis control and elimination 2010-50: cure, care, and social development. Lancet 2010; 375 (9728): 1814–29.

18. Al Yaquobi F, Al-Abri S, Al-Abri B, Al-Abaidani I, Al-Jardani A, D’Ambrosio L et al. Tuberculosis elimination: a dream or a reality? The case of Oman. Eur Respir J 2018; 51(1):1–3.

19. Voniatis C, Migliori GB, Voniatis, Georgiou A, D’Ambrosio L, Rosella Centis and Mario C. Raviglione. Tuberculosis elimination: dream or reality? The case of Cyprus. Eur Respir J 2014; 44(2):540–3.

20. Migliori GB, Raviglione MC, editors. Essential Tuberculosis. Cham: Springer International Publishing; 2021.

21. Lienhardt C, Lönnroth K, Menzies D, Balasegaram M, Chakaya J, Cobelens F et al. Translational Research for Tuberculosis Elimination: Priorities, Challenges, and Actions. PLoS Med 2016; 13(3):e1001965.

22. Glaziou P, Falzon D, Floyd K, Raviglione M. Global epidemiology of tuberculosis. Semin Respir Crit Care Med 2013; 34(1):3–16.

23. Duarte R, Migliori GB, Zumla A, Cordeiro CR. Strengthening tuberculosis control to advance towards elimination: The 2018 Rev. Port. Pneumol. (RPP) TB Series. Pulmonology 2018; 24(2):67–8.

24. Goosby E, Jamison D, Swaminathan S, Reid M, Zuccala E. The Lancet Commission on tuberculosis: building a tuberculosis-free world. Lancet 2018; 391(10126):1132–3.

25. WHO. Global Strategy for Tuberculosis Research: RESEARCH AND INNOVATION; 2020.

26. Harris RC, Sumner T, Knight GM, Zhang H, White RG. Potential impact of tuberculosis vaccines in China, South Africa, and India. Sci Transl Med 2020; 12(564).

27. Schrager LK, Vekemens J, Drager N, Lewinsohn DM, Olesen OF. The status of tuberculosis vaccine development. Lancet Infect Dis 2020; 20(3):e28–e37.

28. Georghiou L, Harper JC, Miles I, Kennan M, Popper R, editors. The handbook of Technology Foresight: Concepts and Pratice. Northampton MA USA: Edward Elgar Pub; 2009.

29. Cabral, B. P, Fonseca M, Mota F. What is the future of cancer care? A technology foresight assessment of experts’ expectations. Economics of Innovation and New Technology 2019; 28.

30. Haghdoost A, Pourhosseini SS, Emami M, Dehnavieh R, Barfeh T, Mehrolhassani MH. Foresight in health sciences using CLA method. Med J Islam Repub Iran 2017; 31:1–8.

31. Martin B. R. The origins of the concept of ‘foresight’ in science and technology: An insider’s perspective. Technological Forecasting and Social Change 2010; 77(9):1438–47.

32. Miles I. The development of technology foresight: A review. Technological Forecasting and Social Change 2010; 77(9):1448–56.

33. Zackiewicz M., Salles-Filho S. Technological Foresight: Um instrumento para política científica e tecnológica [Estudos Prospectivos]. PARCERIAS ESTRATÉGICAS 2001; 10 - March 2001:144–61.

34. Ingelstam L. (Long term science policy: a limited vision: Reseacrch Foresight: priority stting in science [Ben Martin and John Irvine]. Futures 1991:211–3.

35. Dosi G, Soete L, C.R. Freeman, Nelson RR, editors Technical Change and Economic Theory; 1988.

36. Linstone HA. Three eras of technology foresight. Technovation 2011; 31(2-3):69–76.

37. Zackiewicz M, Sales-Filho S. Estudos Prospectivos: Tecnology Foresight - Um instrumento para política científica e tecnológica Março/2001; (PARCERIAS ESTRATÉGICAS número 10).

38. Zenteno-Cuevas R. Successes and failures in human tuberculosis vaccine development. Expert Opin Biol Ther 2017; 17(12):1481–91.

39. Pai M, Behr MA, Dowdy D, Boehme CC, Ann Ginsberg6 Soumya Swaminathan7, Melvin Spigelman8 Haileyesus Getahun9. Tuberculosis. Nature Reveiws 2016:1–23.

40. Kuhn TS, editor. A Estrutura das revolucoes cientificas 1989. São Paulo: Perspectiva; 1998.

41. Abu-Raddad LJ, Sabatelli L, Achterberg JT, Sugimoto JD, Longini IM, Dye C et al. Epidemiological benefits of more-effective tuberculosis vaccines, drugs, and diagnostics. Proc Natl Acad Sci U S A 2009; 106(33):13980–5.

42. Gupta S, Kakkar V. Recent technological advancements in tuberculosis diagnostics - A review. Biosens Bioelectron 2018; 115:14–29.

43. Schrager kl, Harris RC, Vekemans j12. Research and development of new tuberculosis vaccines: a review. F1000RESEARCH 2018.

44. Bonaccorsi A, Apreda R, Fantoni G. Expert biases in technology foresight. Why they are a problem and how to mitigate them. Technological Forecasting and Social Change 2020; 151:1–17.

45. Bolger F, Wright G. Use of expert knowledge to anticipate the future: Issues, analysis and directions. International Journal of Forecasting 2017; 33(1):230–43.

46. MacGregor-Fairlie M, Wilkinson S, Besra GS, Goldberg Oppenheimer P. Tuberculosis diagnostics: overcoming ancient challenges with modern solutions. Emerg Top Life Sci 2020; 4(4):423–36.

47. Working Group on New TB Drugs. Global TB new drugs pipeline. Stop TB partnership; 2022. Available from: URL: https://www.newtbdrugs.org/pipeline/clinical.

48. Lundvall B-Å. National Innovation Systems—Analytical Concept and Development Tool. Industry and Innovation 2007; 14(1):95–119.

49. Edquist C. The Systems of Innovation Approach and Innovation Policy: An account of the state of the art. Aalborg; 2001. Available from: URL: http://www.tema.liu.se/tema-t/sirp/chaed.htm.

50. Dosi G. Technological paradigms and technological trajectories: A suggested interpretation of determinants and directions of technical change. revista brasileira de inovação 1982:17–32.

51. Rosemberg N. Quão Exógena é a ciência? revista brasileira de inovação 2006; 2006/5(2):245–271.

52. HUSSEY G. Pipeline Report 2021: Tuberculosis vaccines.

53. Li JF, Garnsey E. Policy-driven ecosystems for new vaccine development. Technovation 2014; 34(12):762–72.

54. Kaufmann SHE. Learning from natural infection for rational tuberculosis vaccine design:From basic science to trans 2010.

55. Kaufmann SHE. Future vaccination strategies against tuberculosis: thinking outside the box. Immunity 2010; 33(4):567–77.

56. WHO. consolidated guidelines on tuberculosis: Rapid diagnostics for tuberculosis detection; Module 3: Diagnosis. Geneva, Switzerland: World Health Organization.

57. WHO. consolidated guidelines on tuberculosis: Rapid diagnostics for tuberculosis detection; Module 3: Diagnosis. Geneva, Switzerland: World Health Organization.

58. Miller BS, Gliddon HD, McKendry RA. Towards a Future of Rapid, Low-Cost, Multiplexed Detection of Antimicrobial Resistance Markers for Tuberculosis and Other Pathogens. Clin Chem 2019; 65(3):367–9.

59. WHO. Rapid communication: TB antigen-based skin tests for the diagnosis of TB infection. WHO; 2022.

60. Darboe F, Mbandi S, Thompson EG, Fisher M, Rodo M, van Rooyen M et al. Diagnostic performance of an optimized transcriptomic signature of risk of tuberculosis in cryopreserved peripheral blood mononuclear cells. Tuberculosis (Edinb) 2018; 108:124–6.

61. Zak DE, Penn-Nicholson A, Scriba TJ, Thompson E, Suliman S, Amon LM et al. A blood RNA signature for tuberculosis disease risk: a prospective cohort study. The Lancet 2016; 387(10035):2312–22.

62. Levin M, Kaforou M. Predicting active tuberculosis progression by RNA analysis. The Lancet 2016; 387(10035):2302–11. Available from: URL: http://dx.doi.org/10.1016/S0140-6736(16)00165-3.

63. MacLean E, Broger T. A 10-Gene Signature for the Diagnosis and Treatment Monitoring of Active Tuberculosis Using a Molecular Interaction Network Approach. EBioMedicine 2017; 15:112–26.

